# Long-term follow-up in pontocerebellar hypoplasia type 2A: survival, symptom course, and disease burden

**DOI:** 10.64898/2026.09.10.26362700

**Authors:** Alice Kuhn, Anna-Lena Klauser, Johanna Engel, Antonia Herrmann, Maren Hackenberg, Julia Matilainen, Simone Mayer, Saskia Frölich, Ingeborg Krägeloh-Mann, Samuel Groeschel, Wibke G. Janzarik

## Abstract

**Background:** Pontocerebellar hypoplasia Type 2A (PCH2A) is a rare, autosomal recessive disorder with reduced life expectancy. Clinical features include profound developmental delay, microcephaly, and a dyskinetic movement disorder. Additional symptoms frequently observed are feeding difficulties, vomiting, dystonic attacks, and seizures. Compared to parents of typically developing children, parents of children with PCH2A experience decreased health-related quality of life. The aim of this study was to provide a follow-up on survival, symptoms, and perceived disease burden.

**Methods:** Patients were primarily recruited via the German patients’ organization (PCH-Familie e.V.). Inclusion required genetic confirmation of PCH2A. Data were collected from 2020-2022 using parent questionnaires and medical reports; uncertainties were clarified during semi-standardized telephone interviews. Statistical analysis was quantitative, appropriate statistical tests were used, depending on the data distribution.

**Results:** Sixty-five patients participated, with a median age of 6.4 years and balanced gender distribution. Sixteen patients (25%) died at a median age of 5.5 years. Ten-year survival rate was 71.5%. Feeding difficulties, dyskinesia, and sleep disturbances were common and persistent symptoms. Seizures increased with age, whereas vomiting and dystonic attacks decreased over time. Perceived disease burden differed among families, but most reported overall impaired quality of life. The most burdensome symptoms were restlessness, sleep disorder, and gastrointestinal issues, while seizures/epilepsy were ranked as least impactful. With increasing age, perceived burden from feeding and swallowing disorders, gastrointestinal problems, episodic abdominal pain, and overall quality of life decreased significantly.

**Conclusions:** Life expectancy in PCH2A remains limited, but survival appears to have improved in more recent birth cohorts. Most symptoms persist with age, though a few, like reflux and dystonic attacks, improve in tendency. Conversely, respiratory symptoms and seizures tend to worsen. Notably, perceived disease burden generally decreases as patients age. Restlessness proved to be the most burdensome symptom, although it has not been clearly described before.

## Background

Pontocerebellar hypoplasia type 2 (PCH2A) is a rare, autosomal recessive neurodevelopmental disorder resulting from a homozygous pathogenic variant in the TSEN54 gene (p.A307S) [1]. The clinical phenotype is marked by severe developmental delay, microcephaly, failure to thrive, and multi-organ involvement, reflecting significant impact on the nervous and other systems [2]. The first systematic Natural History Study (NHS) on PCH2A was conducted and published by the Department of Child Neurology at the University of Tübingen in 2014 [2]. This foundational study established important aspects of the clinical course and expanded the known symptom spectrum [2].Throughout this manuscript, the initial study is referenced as the first NHS, while the current work is termed the second NHS—serving as an updated and expanded follow-up. Although it is well known that PCH2A significantly shortens life expectancy, isolated reports document survival into adulthood [2,3]. Precise estimation of long-term survival, however, remains unclear. The only available survival analysis to date originates from the first NHS, which included only those patients who reached the age of ten years or who had died younger [2]. In recent years, there have been advances in symptomatic management, even though a causal therapy remains unavailable. One of the core questions of this second NHS, therefore, is whether these advances have led to improved survival outcomes.

The causes of death in PCH2A have not been thoroughly characterized. While the first NHS often found the cause of death unclear, additional literature suggests that respiratory complications such as pneumonia and apnea are frequently involved—not only in PCH2A but also in other severe neuropediatric disorders [2,4,5]. This study aims to gain a clearer understanding of the causes of death in PCH2A, thereby enabling improved clinical management and more informed counselling for families.

Symptom progression and variability in PCH2A remain insufficiently described, particularly over the extended lifespan now observed in some patients. In the neonatal period, common features include feeding difficulties, abnormal muscle tone, jitteriness, hypersomnolence, and respiratory issues [2]. Soon after, hallmark dyskinetic movement disorders (predominantly choreoathetosis), sleep disturbances, and excessive vomiting typically emerge [2]. Other features—including seizures, dystonic attacks, infections, and disturbed temperature regulation—often develop later in childhood [2]. MRI findings not only reveal pontocerebellar hypoplasia but also indicate concurrent cortical atrophy, supporting the hypothesis of progressive neurodegeneration [6,7].

Sánchez et al. were among the first to more specifically describe PCH2A in a larger cohort, including the identification of dystonic attacks as a core symptom [2]. Janzarik et al. reported for the first time gastrointestinal manifestations such as meteorism, constipation, and episodic abdominal pain in several patients [8]. As patients survive longer due to improved care, it becomes increasingly important to map the temporal sequence and long-term evolution of both common and newly recognized symptoms, providing a solid base for family counselling and management.

Quality of life for families is profoundly affected [9]. Previous research specifically documents a reduction in quality of life among mothers with a child suffering from PCH2A [9]. However, to date, little is known about which individual symptoms are perceived as most debilitating, or how the burden evolves over time. Data on the age-related progression of symptom burden and quality of life remain scarce.

In summary, we are therefore the first to be able to chart longitudinal trends over a period of around a decade, right through to adulthood, in comparison with the fist NHS. Accordingly, the primary objectives of this enlarged, updated natural history study were threefold: To examine whether survival has improved in PCH2A, to elucidate the progression, sequence, and transformation of symptoms across the lifespan, and to assess which symptoms most contribute to disease burden and diminished quality of life, with special attention to changes as patients age. This study seeks to fill notable knowledge gaps, providing comprehensive data that can support more targeted clinical intervention and more informed guidance for affected families.

## Methods

The results presented in this manuscript are derived from the second NHS on PCH2A, conducted between December 2020 and September 2022 utilizing a detailed parent questionnaire.

This manuscript focuses on survival, the range and progression of symptoms over time, and overall symptom burden. Additional outcomes from the second NHS with different thematic focuses have been published separately and are referenced where relevant [7,10]. The study received ethical approval from the University of Freiburg in November 2020 (No. 20–1040), as well as from the University of Tübingen in 2012 (No. 105/2012BO2) and 2021 (No. 961/2020BO2). It is registered in the German Clinical Trials Register (“Deutsches Register Klinischer Studien”, ID: DRKS00022511).

Data were collected from a cohort of patients with genetically confirmed PCH2A, primarily recruited via the German patient network PCH-Familie e.V. Written informed consent was obtained from legal guardians for all patients. In instances where genetic confirmation was unavailable, inclusion was based on consistent phenotypic presentation and genetic confirmation in a sibling. Both living and deceased patients were included. The study population aligns with that reported by Kuhn et al. and Herrmann et al., and overlaps partially with Pretzel et al.[7,10,11]. The entire cohort was divided into subgroups: approximately half of the patients’ data was acquired from 2020-2022 (second NHS), while a smaller subset was collected in 2012 (first NHS, [2]), with unknown survival status for those alive in 2012. A longitudinal cohort was generated from patients who participated in both studies.

Data management and collection were executed using REDCap electronic data capture tools hosted at the University of Freiburg [12] and Microsoft Excel (Microsoft Corporation. Microsoft Excel for Windows, Microsoft® Excel® LTSC Professional Plus 2024, Version 2408). Questionnaire responses were validated against medical records, and remaining discrepancies were resolved via final telephone interviews with parents. Additionally, data from the first NHS were analyzed for those who participated only in 2012 but not in the follow-up (n=12) [2]. Symptom presence or persistence was assessed relative to the time of each survey. Thus, for patients in the first NHS only, survey data refer to 2012, contingent on whether the symptom was covered in the earlier questionnaire. Due to differences in questionnaire scope, missing data were more common for patients who only participated in the first NHS. Point prevalence measurement also varied: the second NHS referred to the year prior to survey, whereas timing was less defined in the first NHS.

SPSS v29.0.0.0 (IBM Corp., Armonk, NY, USA) and R (R Foundation for Statistical Computing, Vienna, Austria) were used for statistical and graphical analysis. Descriptive statistics (means, medians, interquartile range (IQR), and percentages, as appropriate) were calculated to describe the characteristics of patients, causes of death, the symptom overview (including age-dependent and long-term follow-up) such as the disease burden. Kaplan-Meier-Estimate with right censoring including the 95% confidence interval (95% CI) was used to assess survival.

Two patients died shortly after data acquisition and were included as deceased into the survival analysis, but clinical data was collected while the patients were still alive. One of those patients was part of the follow-up cohort, resulting in 13 patients being alive at the end of the study was reported in table 1, and 14 patients contributing to the longitudinal follow-up in Figure 4. The family of one patient who participated in the first NHS provided the patient’s current age but no further clinical data for the second NHS: regarding survival the age at the time of the second NHS is included whereas regarding symptom course over time the age at the last follow up is used.

Fishers exact test was used for comparison of survival at ten years of age (whereas for overall survival, Log-rank test was used) and for comparison of symptoms over age due to small group samples and unequally distributed frequencies. To adjust for multiple testing, a post-hoc analysis using Benjamin-Hochberg for FDR control was performed subsequently. For scaled data, either ANOVA followed by pairwise t-tests with Bonferroni post-hoc correction, or Kruskal-Wallis test followed by pairwise Wilcoxon rank tests with Bonferroni post-hoc correction, was applied depending on data distribution.

Perceived disease burden for each symptom was assessed using a visual analogue scale (VAS) from 0 (no burden) to 10 (very high burden). This is not a validated (health-related) quality of life instrument, but rather a parent-related measure of impaired quality of life, and is therefore referred to as the ‘perceived disease burden’. Median scores were visualized using heat maps for both patients and parents.

## Results

### Study population

This study analyzed data from 65 patients with genetically confirmed PCH2A (see Table 1). Of these, 32 individuals were newly recruited and had not participated in the first Natural History Study (NHS) conducted in 2012. Twenty-one patients (from a total of 33 included in the first NHS) took part in both studies [2]. The mean age at time of death (mean for better comparison with the first NHS) was 9 years. The longitudinal disease course was assessed using data from these 21 patients. In 2022, the median age of this subgroup was 15.1 years, and gender distribution was balanced (Table 1).

**Table 1.** Study population:

|  | Entire cohort | Longitudinal cohort |
| --- | --- | --- |
| **Number of patients (%)** | 65/65 (100%) | 21/65 (32.3%) |
| Median age in years<sup>1</sup> (IQR) | 6.4 (11) | 15.1 (8) |
| **Alive (%)** | 42/65 (64.6%) | 13/21 (61.9%) |
| Median age in years<sup>1</sup> (IQR) | 5.5 (12) | 16.0 (2) |
| **Deceased (%)** | 16/65 (24.6%) | 8/21 (38.1%) |
| Median age in years<sup>1</sup> (IQR) | 7.2 (8) | 6.4 (11) |
| **Gender** |  |  |
| Female (%) | 33/65 (50.8%) | 10/21 (47.6%) |
| Male (%) | 32/65 (49.2%) | 11/21 (52.4%) |

<sup>1</sup>at last follow-up

### Survival

In the second NHS, 42 patients were alive, 16 had died, and the survival status of 7 patients was unknown (Table 1, Figure 1a). Among the 33 patients in the first NHS cohort, 24 were alive at the time of the initial study [2]. Of these, 14 (58.3%) were still alive in the second NHS, with clinical follow-up data available for 13; three patients (12.5%) died in the interim (including one shortly after data acquisition in the second NHS). For the remaining 7 of 24 patients (29.2%), survival status could not be determined due to loss to follow-up.

Marked variability in outcomes was observed: five patients died before the age of five, whereas the oldest surviving patient was 33 years at the last follow-up (Figure 1a). The five-year survival rate for the entire cohort was 92.0% (95% CI: 85-99%), although a considerable proportion of patients were censored at an early age (Figure 1b).

**Figure 1.**
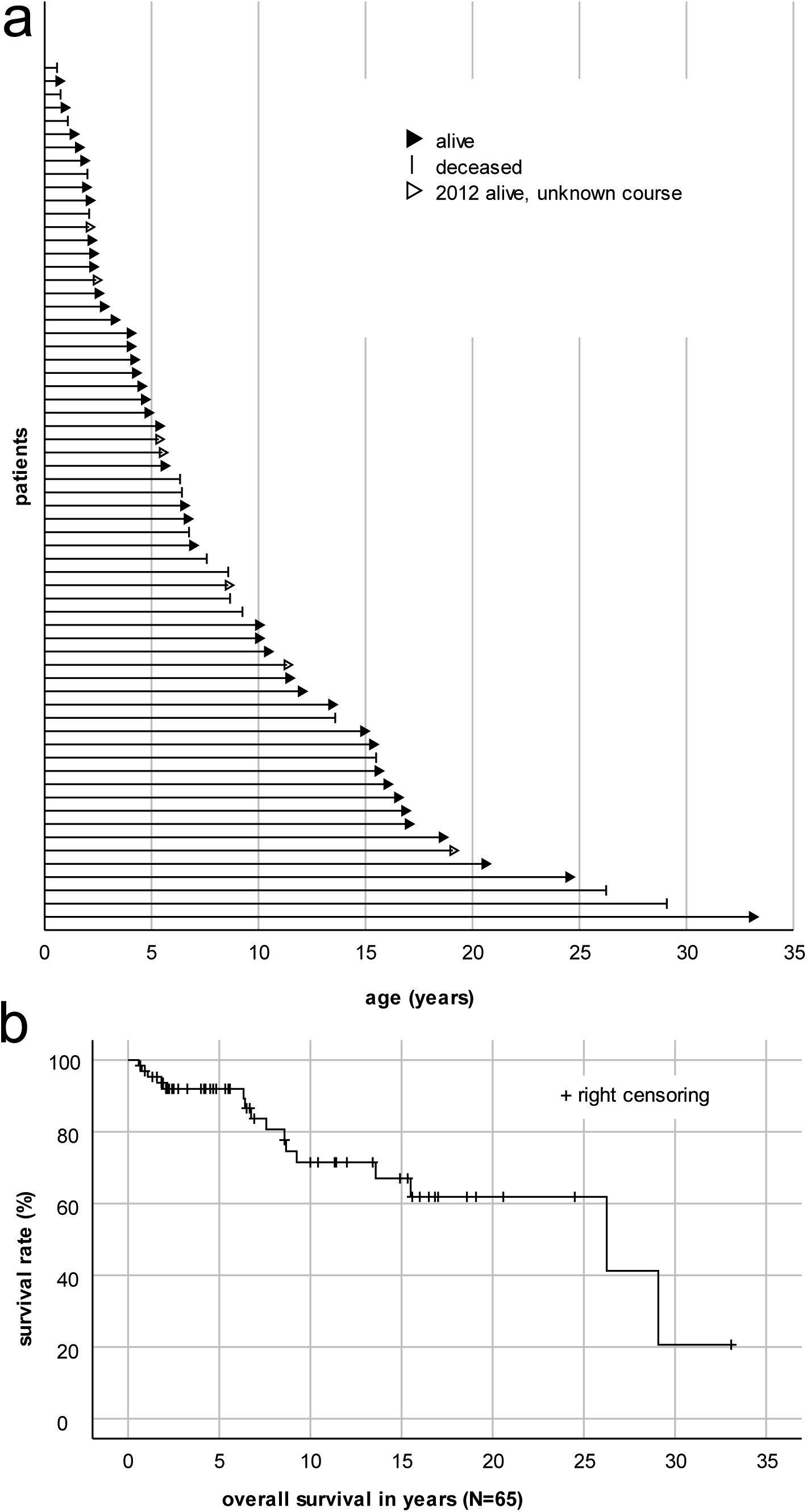
Overall survival: a) At the time of the second NHS, 42 patients were alive, 16 were deceased, and the survival status of 7 patients was unknown due to loss to follow-up. b) Overall survival for the entire cohort was estimated using the Kaplan–Meier-Estimate with right censoring (modified from Kuhn et al. [10], with minor changes in the lettering).

**Figure 2.**
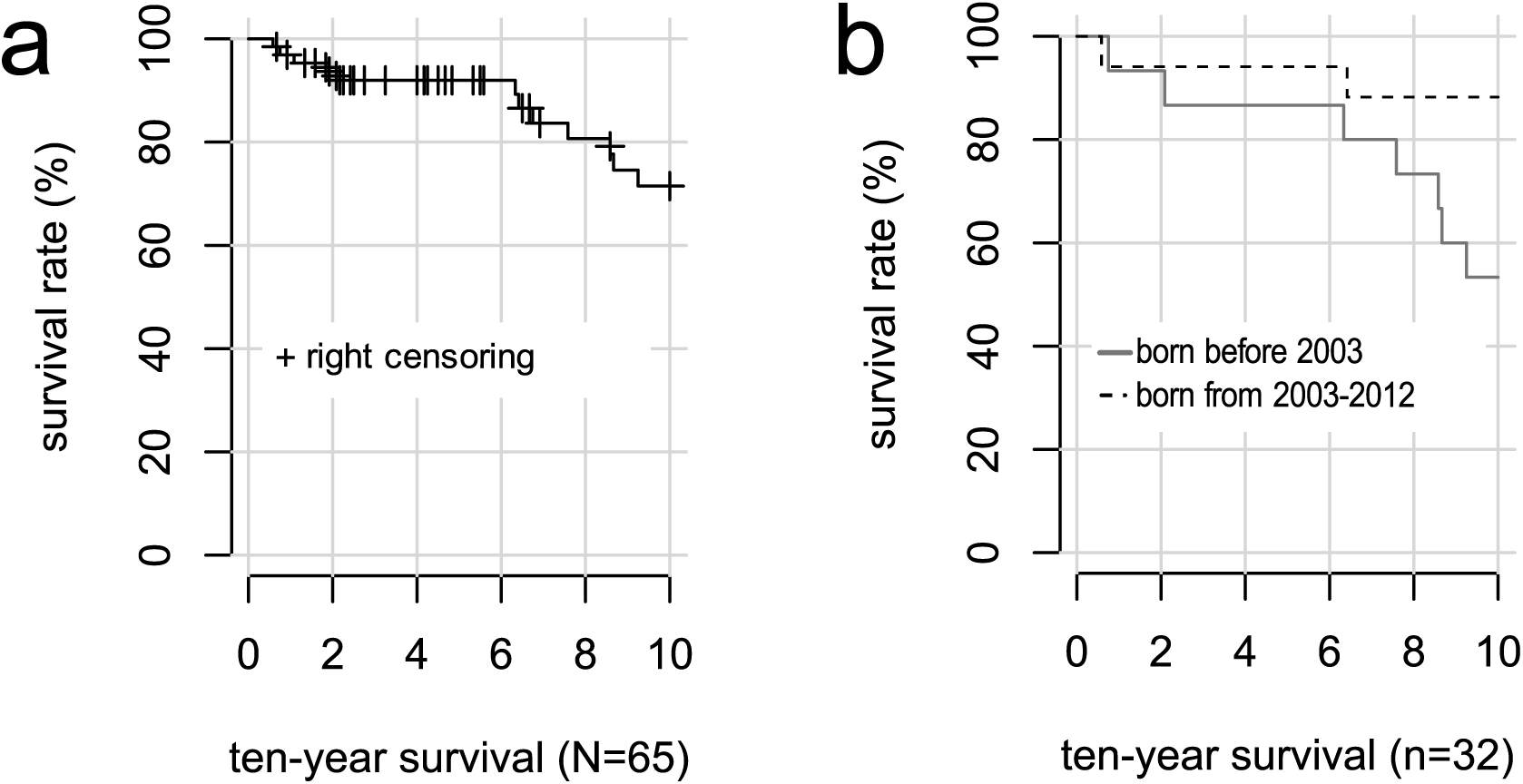
Ten-year survival: a) Ten-year survival of the total cohort (N=65) estimated using the Kaplan-Meier-Estimate with right censoring, censored observations: n = 30 (two patients at the exact age of ten years excluded). b) Ten-year survival was further evaluated in the subgroup of patients who either reached ten years of age or died before ten, comparing subcohorts born before 2003 (n=15) and from 2003 to 2012 (n=17).

The ten-year survival rate for the full cohort was 71.5% (95% CI: 58-88%), with 30 patients censored as they were younger than ten years at last follow-up, and 12 patients having died before reaching ten years of age (Figure 2a). Among patients born before 2003, the ten-year survival rate was 53.3% (95% CI: 33-86%), calculated only for those who had either attained ten years of age by last follow-up or died earlier. For patients born between 2003 and 2012, the corresponding ten-year survival rate was 88.2% (95% CI: 74-100%) (Figure 2b). Ten-year survival was significantly higher in the latter group (p=0.049). However, comparison of overall survival between these two subgroups did not yield a statistically significant difference. Parents were asked to provide the causes of death for their children in free-text format. These responses were subsequently categorised as follows: gradual decline (4/16), aspiration (4/16), pneumonia (2/16), respiratory arrest (2/16), organ failure (1/16), and unknown causes (3/16).

### Clinical Symptoms

Clinical symptoms in PCH2A can be grouped into gastrointestinal, respiratory, movement disorders, seizures, and other categories (Figure 3). Nearly all patients experienced feeding difficulties, dyskinesia, and sleep disturbances. Severe episodic restlessness and reflux were also reported in almost all cases where families provided information. In some patients, seizure occurrence could not be clearly determined, as distinguishing seizures from the movement disorder proved challenging.

**Figure 3.**
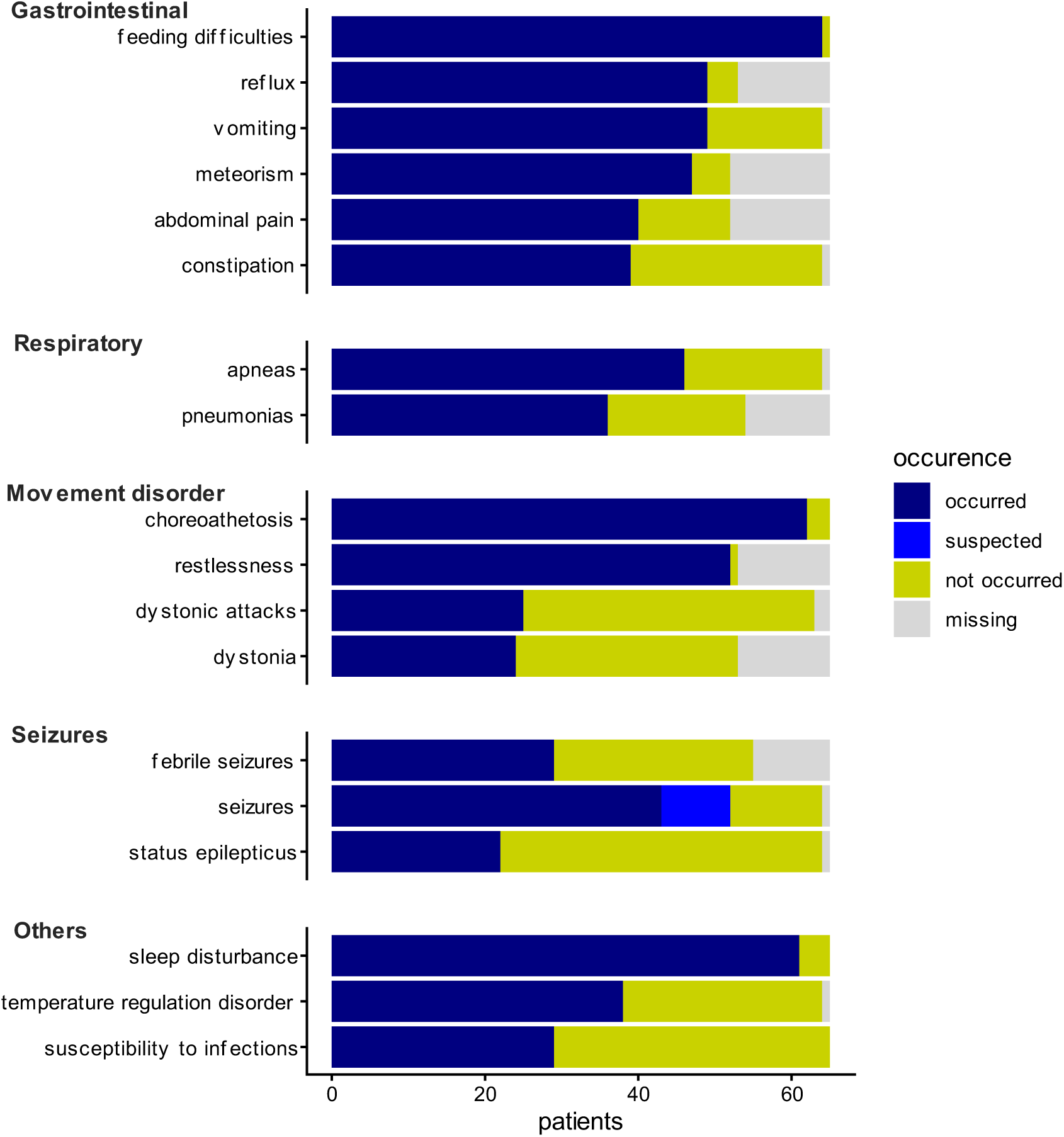
Life time prevalence of clinical symptoms. For all symptoms, except for seizures, parents were asked if a symptom occurred or not. For seizures, suspected occurrence was asked for only in the second NHS.

To compare characteristic clinical symptoms of patients with PCH2A over time, point prevalence was evaluated across three age groups at last follow-up: under 2 years (10 patients), 2–9 years (33 patients), and over 10 years (22 patients). All surviving patients from the first NHS [2] were assigned to the oldest group. For most symptoms, prevalence at last follow-up was assessed, while for pneumonia, status epilepticus, and susceptibility to infections, lifetime prevalence was analysed, as point prevalence was considered inappropriate (Table 2). Feeding difficulties, meteorism, choreoathetosis, restlessness, and sleep disturbances were highly prevalent across all age groups. Reflux, abdominal pain, and—tendentially—dystonia were less common in older patients. The prevalence of vomiting, constipation, dystonic attacks, and susceptibility to infections appeared to increase in younger age groups before declining again. In contrast, respiratory symptoms, seizures, and difficulties with temperature regulation tended to become more frequent with advancing age (Table 2).

**Table 2.**
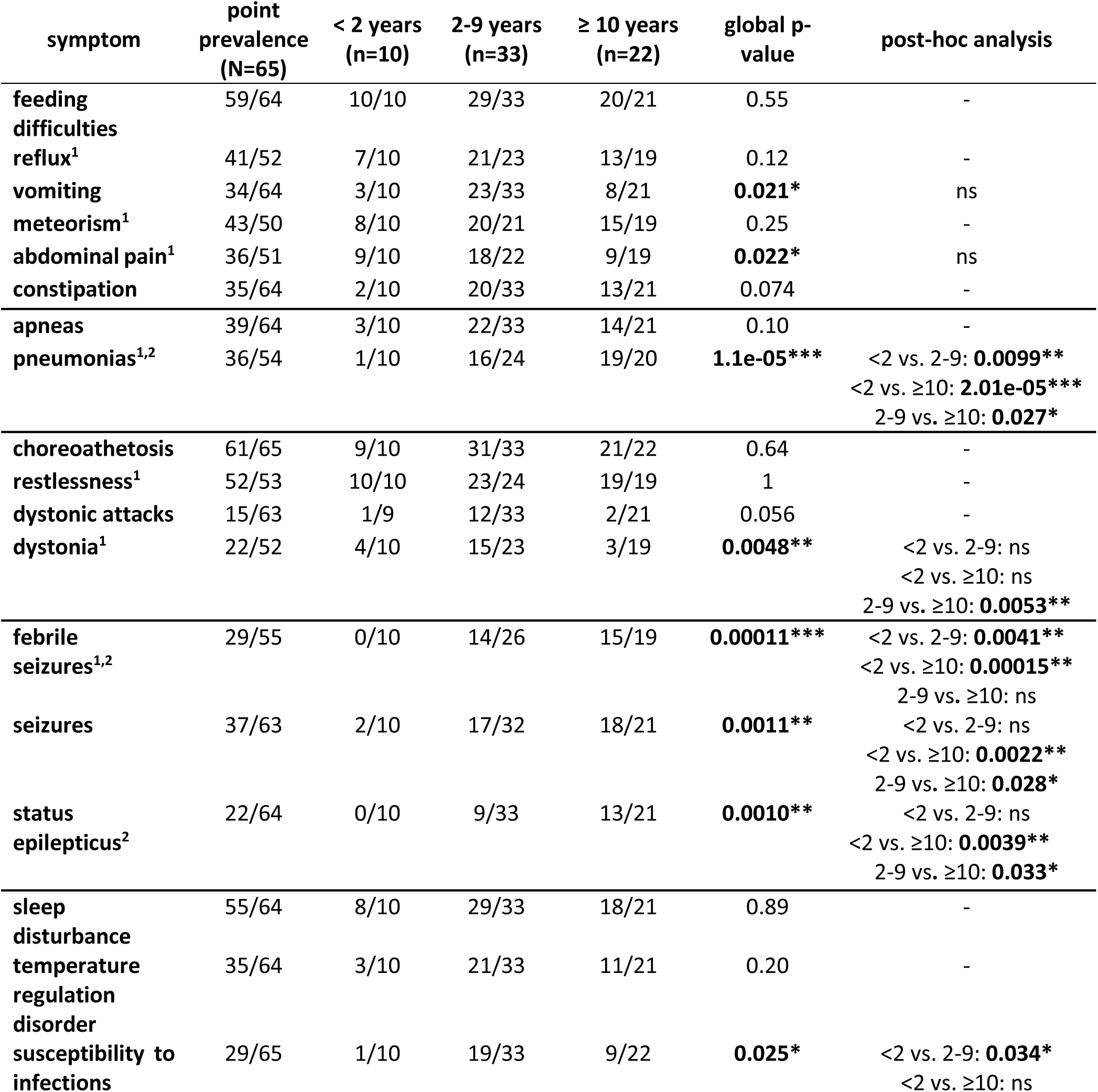
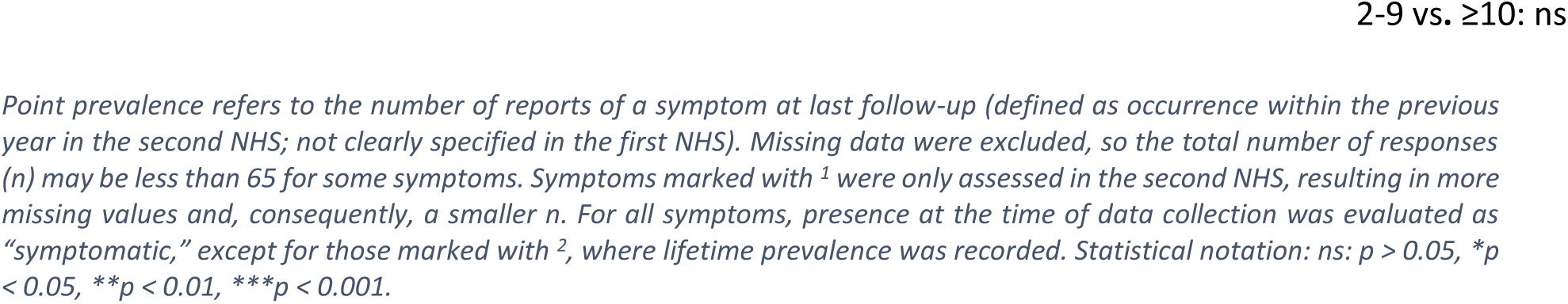
Point prevalence of clinical symptoms, broken down by age group.

These findings were largely consistent with the longitudinal data, except for susceptibility to infections and sleep disturbances, which both exhibited an increasing trend with age (Figure 4).

**Figure 4.**
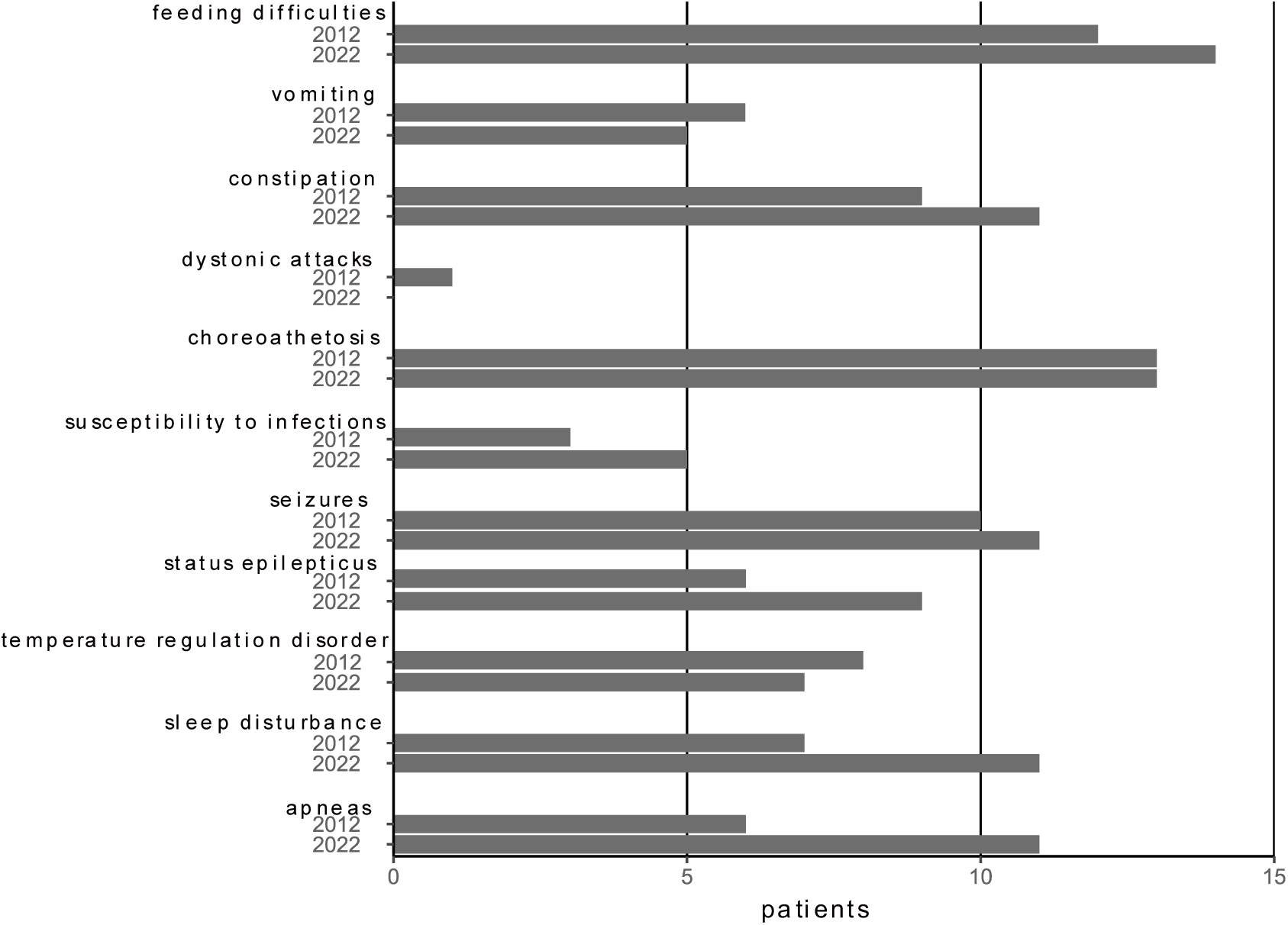
Longitudinal follow-up was available for 14 patients who participated in both data collections (first and second NHS; all were alive at the time of data acquisition, with one patient passing away shortly thereafter). For these individuals, all symptoms assessed in both studies and their evolution over time are depicted, categorising symptoms as present at the first NHS and still present at the second NHS.

### Perceived disease burden

In the 2022 survey, parents evaluated the overall impact of PCH2A on the quality of life for both their affected child and themselves, as well as the burden imposed by frequent clinical symptoms. Parents reported a marked impairment in quality of life, with median scores of 7.7 for their child and 8.2 for themselves. Restlessness (child: 7.2; parent: 7.5), episodic abdominal pain (child: 6.4; parent: 6.0), and sleep disturbances (child: 6.0; parent: 7.1) were identified as the most burdensome symptoms. In contrast, seizures and epilepsy were associated with the lowest reported burden (child: 1.5; parent: 1.3) (Figure 5).

**Figure 5.**
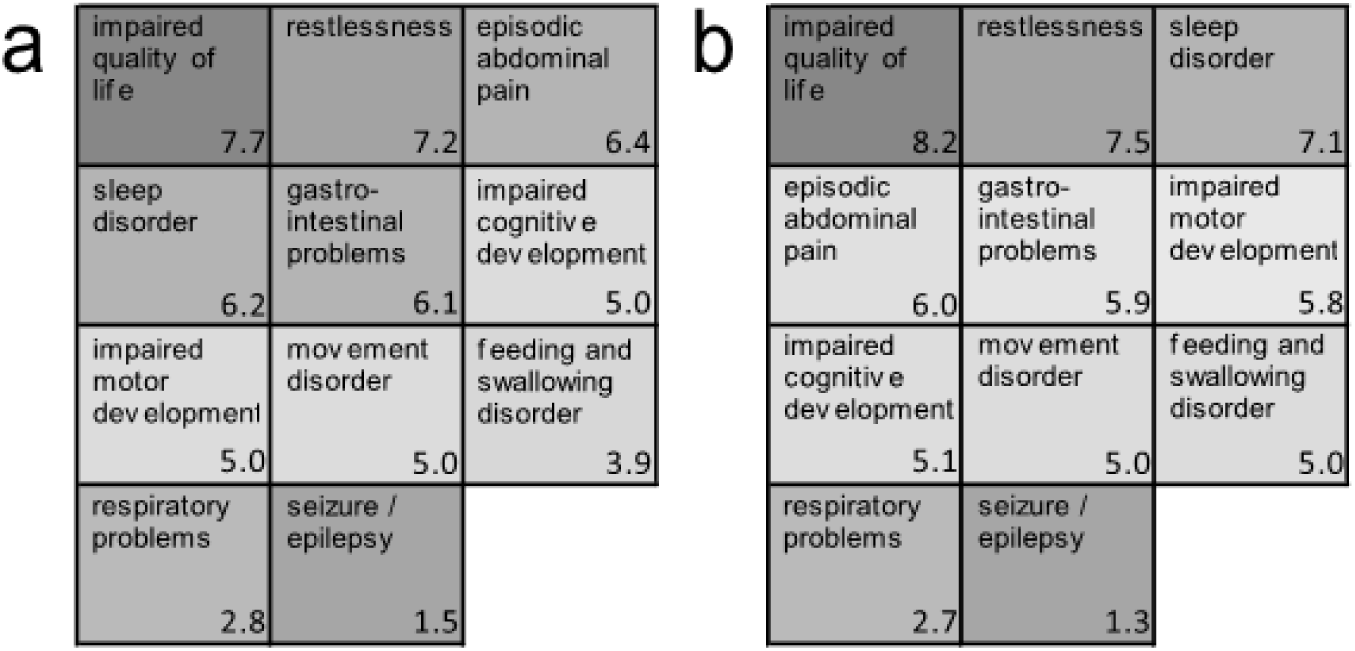
Perceived disease burden in PCH2A patients and self: a) children; b) parents/caregivers; Rating scale ranged from 0 = no impairment/no disease burden, to 10 = very high impairment/disease burden.

Overall impairment in quality of life and perceived disease burden were assessed across three age groups: under 2 years (n = 10), 2–9 years (n = 33), and over 10 years (n = 22) (Figure 6). The oldest group reported a significantly lower degree of impaired quality of life for both children (median 5.0, p= 0.0097) and parents (median 5.9, p = 0.038). Perceived disease burden from gastrointestinal symptoms—including general gastrointestinal complaints and episodic abdominal pain—was also reduced in this group, and parents indicated fewer feeding difficulties (all p < 0.05). In contrast, burden related to restlessness, movement disorders, sleep disturbances, and impaired cognitive or motor development remained stable across age groups. Respiratory symptoms tended to increase, and seizures/epilepsy exhibited a slight age-associated trend; however, neither reached statistical significance.

**Figure 6.**
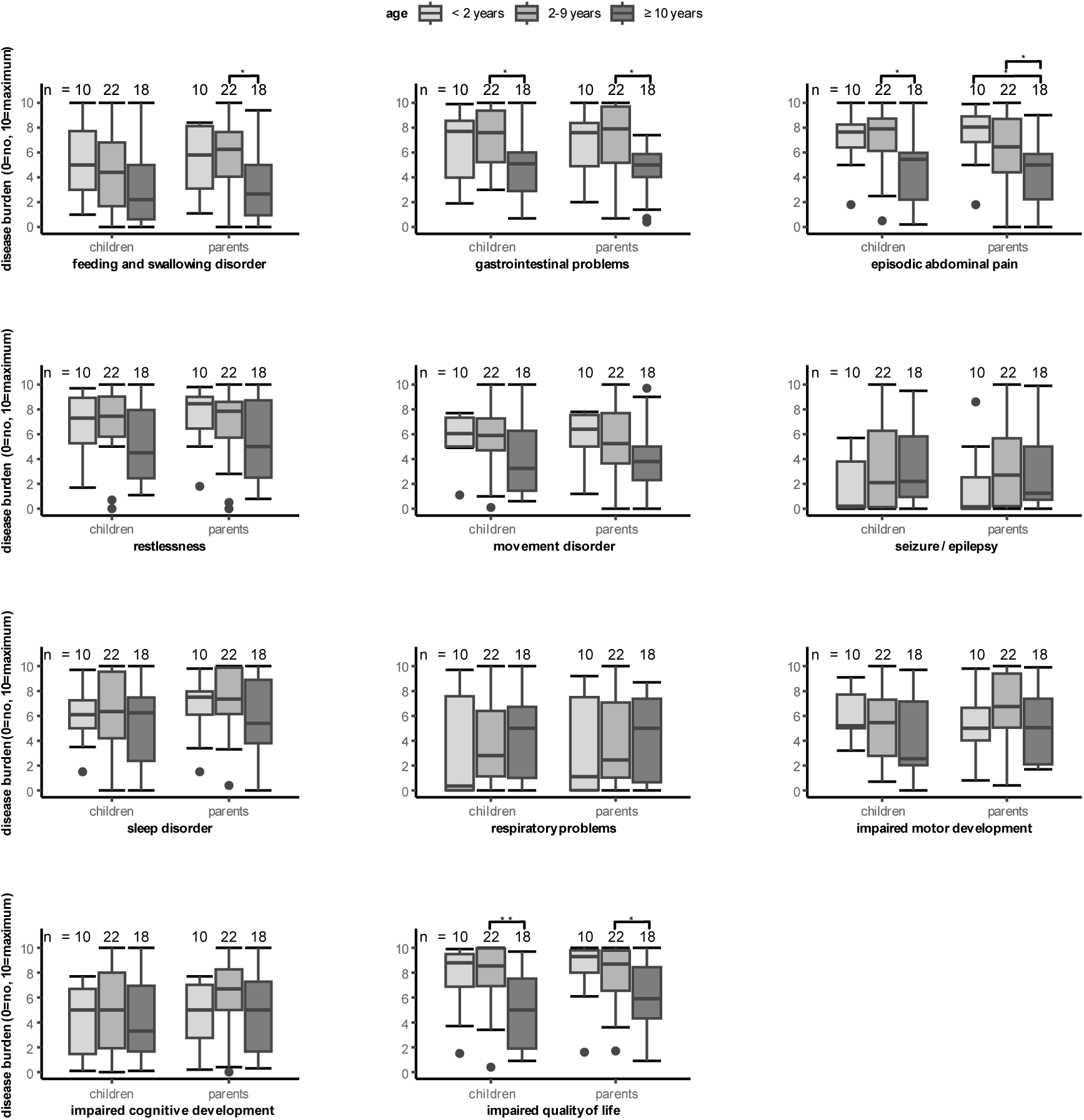
Age-dependent perceived burden of key symptoms in PCH2A: Boxplots illustrate parental ratings of perceived disease burden for specific symptoms, as well as overall quality of life impairment in patients with PCH2A, across three age groups. Ratings range from 0 (no impairment or disease burden) to 10 (very high impairment or disease burden). Statistical significance is indicated as follows: *p < 0.05, **p < 0.01.

## Discussion

The objective of this study was to examine survival, the clinical course of symptoms over time, and the perceived disease burden in PCH2A as part of a comprehensive natural history study. Although patients with PCH2A continue to face reduced life expectancy, as previously described in the literature [2,3], our findings suggest a trend toward improved survival over recent years. The mean age at death in our cohort was nine years, notably higher than the 6 years and 7 months reported in the first NHS [2]. The ten-year survival rate for the entire cohort was 71.5%, compared to approximately 50% reported in 2012 [2]. Patients born between 2003 and 2012 exhibited significantly higher ten-year survival than those born before 2003, and only a few individuals of the longitudinal cohort died between the two NHS assessments. To our knowledge, apart from the study by Sánchez et al., no additional survival analyses for PCH2A have been published. The observed increase in survival may be related to advances in overall care, including medical progress, improved socio-medical and nursing support, as well as symptomatic therapies. These observations warrant further investigation and are especially important for family counselling. Future studies should focus on identifying factors influencing survival. Additionally, increasing survival into adulthood highlights the need to address the transition to adult care.

Respiratory complications, such as aspiration and pneumonia, are major life-threatening risks in patients with PCH2A. In this cohort, respiratory causes—including aspiration, respiratory arrest, and pneumonia—were the most frequent causes of death among deceased patients. These results are in line with previous reports and are characteristic of individuals with complex neurological disorders [2,3,5]. Respiratory infections are a major contributor to morbidity and mortality in this population. It is important that caregivers are made aware of these risks and that preventive strategies are prioritized. Recommended measures include seasonal vaccinations, access to supportive equipment such as pulse oximeters, suction devices, and cough assist devices, as well as care provided by qualified nursing staff [13].

Patients with PCH2A exhibit symptoms affecting multiple organ systems. Most symptoms appear to remain stable over time, with no clear evidence of substantial change. Feeding difficulties, meteorism, choreoathetosis, sleep disturbances, and restlessness are highly prevalent across all age groups, while complete resolution of clinical symptoms is rare. These findings are consistent with earlier studies on PCH2A [2,3]. In the present study, gastrointestinal symptoms—including episodic abdominal pain and meteorism—were assessed in greater detail, now described in a larger cohort, whereas these symptoms had previously been reported only in case reports [8]. Additionally, restlessness and dystonia, alongside choreoathetosis and dystonic attacks, were further characterized. Notably, gastroesophageal reflux was among the few symptoms that showed a decrease with age, which may be attributable to frequent use of proton pump inhibitors (PPIs) and surgical treatments such as PEG placement and fundoplication [10].

In contrast, susceptibility to infections was observed to increase initially and then decrease with age. This pattern is also seen in typically developing children and is frequently attributed to increased exposure in external childcare environments, such as daycare or kindergarten settings [14]. Additionally, the term “susceptibility to infections” may be somewhat misleading, as based on current evidence does, there is no indication that *TSEN54* loss-of-function impairs immune function. Furthermore, the peak in so-called susceptibility to infection tends to suggest a high incidence of infections during the typical risk period of 2–9 years of age. Instead, patients with PCH2A commonly experience prolonged or more severe infections due to impaired clearance of respiratory secretions, a consequence of severe disease and pronounced motor impairment [5]. This mechanism may also account for the observed rise in pneumonia with advancing age, as cumulative microaspirations can lead to clinically manifest pneumonia[15].

The observed increase in seizures, status epilepticus, and apnea may indicate progressive cerebral involvement, as supported by evidence of progressive atrophy [6,7]. These findings are largely confirmed by the longitudinal cohort. To date, there are no published data on the efficacy of medications in PCH2A. Further clinical studies are needed to clarify the impact of pharmacological interventions on symptom evolution throughout the disease course. Moreover, the underlying pathophysiological mechanisms remain incompletely understood—particularly regarding whether symptoms result exclusively from the neurological disorder itself or if they may also reflect direct effects of the *TSEN54* variant on different organs.

Restlessness is rated as the most burdensome symptom, concordant with the fact that almost all patients suffer from severe restlessness episodes and that restlessness persists over age. The high prevalence of seizures contrasts with the relatively low symptom burden attributed to seizures/epilepsy, followed by respiratory problems. At first glance, this may seem paradoxical, as epileptic seizures are typically regarded as highly distressing in otherwise healthy individuals [16]. However, there is limited data on how epilepsy specifically impacts quality of life in children with underlying neurological conditions [17]. Since respiratory causes are the most common cause of death in PCH2A, it is also surprising that the associated symptom burden is rated as low. One possible explanation is that a substantial proportion of patients had not yet experienced seizures or respiratory problems at the time of the survey, as these symptoms tend to occur more frequently with increasing age, whereas restlessness starts, occurring early in life.

Total symptom burden, as well as the burden of most individual symptoms, decreased over time, although the symptoms themselves generally persisted. In contrast, the burden of epilepsy and respiratory problems increased, consistent with their clinical trajectory. Several factors may explain the overall reduction in perceived burden. One possibility is a habituation effect, whereby symptoms are perceived as less distressing over time [18]. Alternatively, survival bias may play a role: patients with more severe disease may die earlier, resulting in a cohort enriched for individuals with milder phenotypes. Additionally, as children age, families may benefit from external childcare support, which can further alleviate the perceived burden.

Overall, ratings for both children and parents were similar across most items, likely due to the use of parent proxy assessments; children were unable to self-report. Parental burden is also closely influenced by the child’s burden, suggesting a potential bidirectional relationship [19].

Since PCH2A remains an incurable disease associated with a high symptom burden [9], the systematic assessment of symptoms is essential to facilitate the prioritization of therapeutic strategies and supportive care.

### Limitations

A key limitation regarding survival analysis is that a substantial proportion of patients were younger than five years at the last follow-up. Including these individuals with right censoring may lead to an overestimation of survival, whereas excluding them further reduces the sample size and could result in an underestimation of survival.

Direct comparison with the first NHS is also limited, since the cohorts overlap at least in part and the analysis was carried out in various ways (e.g. survival analysis in this publication via Kaplan-Meier-Estimate, whereas in the first NHS only children who reached the age of ten years or died earlier were taken into account to analyse survival).

The reliance on parent-reported questionnaires constitutes an important limitation for the entire study, particularly with respect to determining causes of death in the absence of autopsy data. Furthermore, comparison of symptoms across age groups and evaluation of symptom progression in the longitudinal cohort are restricted by the lack of age stratification in the longitudinal data.

The use of proxy-reported outcomes presents an additional limitation, notably restricting assessment of patient-specific quality of life. Moreover, the symptoms included in the symptom burden assessment did not fully correspond to those assessed for prevalence, representing another methodological limitation.

## Conclusions

This manuscript presents an overview of survival, age-dependent symptoms, and symptom-related perceived disease burden in patients with PCH2A—a severe, incurable neurodevelopmental disorder involving multiple organ systems. Recent data suggest that life expectancy and overall survival have improved compared to earlier reports. Respiratory complications remain the leading life-threatening events. Most symptoms manifest early and persist throughout life; however, epilepsy and respiratory issues become more prevalent with increasing age. The most burdensome symptoms identified are restlessness, episodic abdominal pain, and sleep disturbances. Notably, although most symptoms persist, the perceived symptom burden tends to decrease as patients age.

## Data Availability

The datasets generated and analysed during the current study are not publicly available for data protection reasons but are available from the corresponding author on reasonable request.

## List of abbreviations

PCH2A: Pontocerebellar Hypoplasia Type 2A
NHS: Natural History Study
95% CI: 95% confidence interval
IQR: Interquartile range
VAS: visual analogue scale
PPI: Proton pump inhibitors

## Declarations

### Ethics approval and consent to participate

The study received ethical approval from the University of Freiburg in November 2020 (No. 20–1040), as well as from the University of Tübingen in 2012 (No. 105/2012BO2) and 2021 (No. 961/2020BO2). It is registered in the German Clinical Trials Register (“Deutsches Register Klinischer Studien”, ID: DRKS00022511). Written informed consent was obtained from legal guardians for all patients.

### Declaration of Generative AI

Language refinement was supported by Large Language Models (DeepL, AI Hub). Statistical code was generated with the assistance of ChatGPT, an artificial intelligence language model, and subsequently reviewed and executed by the authors.

### Related publications

Three publications have already been published in peer reviewed journals as part of the second NHS [7,10,11]. A supplementary figure by Kuhn et al. was included with minimal changes [10]. In addition, a doctoral thesis by Kuhn on ‘Gastrointestinal symptoms in pontocerebellar hypoplasia type 2A’ is publicly available. The results of the second NHS were summarised in a leaflet for parents, which is available on the PCH-Familie e.V. website.

### Consent for publication

Written informed consent was obtained from legal guardians for all patients.

## Authors’ contributions

AK: data acquisition, data interpretation, analysis, interpretation, visualization, writing – review and editing, ALK: data acquisition, data interpretation, analysis JE: data interpretation, analysis, writing review and editing, AH: Data acquisition, MH: statistical analysis, JM: Conceptualization, funding acquisition, data acquisition, writing review and editing, SM: Conceptualization, funding acquisition, writing – review and editing, SG: Conceptualization, funding acquisition, data interpretation, writing – review and editing, WJ: Conceptualization, funding acquisition, data acquisition, data interpretation, writing – review and editing, supervision.

## Acknowledgements

SG and IKM are members of the European Reference Network for Rare Neurological Diseases (ERN-RND) – project ID 739510.

